# Digital contact tracing contributes little to COVID-19 outbreak containment

**DOI:** 10.1101/2021.06.21.21259258

**Authors:** A. Burdinski, D. Brockmann, B. F. Maier

**Affiliations:** Institute for Theoretical Biology and Integrated Research Institute for the Life-Sciences, Humboldt University of Berlin, Germany

## Abstract

Digital contact tracing applications have been introduced in many countries to aid in the containment of COVID-19 outbreaks. Initially, enthusiasm was high regarding their implementation as a non-pharmaceutical intervention (NPI). Yet, no country was able to prevent larger outbreaks without falling back to harsher NPIs, and the total effect of digital contact tracing remains elusive. Based on the results of empirical studies and modeling efforts, we show that digital contact tracing apps might have prevented cases on the order of single-digit percentages up until now, at best. We show that this poor impact can be attributed to a combination of low participation rates, a non-flexible reliance on symptom-based testing, low engagement of participants, and delays between testing and test result upload. We find that contact tracing does not change the epidemic threshold and exclusively prevents more cases during the supercritical phase of an epidemic, making it unfit as a tool to prevent outbreaks. Locally clustered contact structures may increase the intervention’s efficacy, but only if the number of contacts per individual is homogeneously distributed, a condition usually not found in contact networks. Our results suggest that policy makers cannot rely on digital contact tracing to contain outbreaks of COVID-19 or similar diseases.

## I. INTRODUCTION

During the ongoing coronavirus disease 2019 (COVID-19) pandemic, the severe acute respiratory syndrome coronavirus type 2 (SARS-CoV-2) infected more than 176 million people and caused more than 3.8 million deaths worldwide up to June 18, 2021 [1]. Among other pivotal measures to mitigate or contain the disease’s spread, the most common one is testing and quarantining of symptomatic individuals [2, 3]. While this intervention is usually rather effective, a substantial proportion of transmissions in the COVID-19 pandemic occur from asymptomatic, paucisymptomatic, or presymptomatic infected individuals, which curbs its success [4–6].

A non-pharmaceutical intervention that can help identifying non-symptomatic, yet infectious individuals is “contact tracing” (CT), where epidemiologically relevant contacts of confirmed index cases are traced and isolated. This procedure effectively shortens the infectious period of potentially infected secondary cases, thereby reducing the number of tertiary infections. However, if the tracing mechanism takes too much time to identify and isolate contacts, few further infections are prevented [7], a problem that many countries face when their public health system is overburdened. With the intention to accelerate and supplement the manual tracing process, digital contact tracing (DigCT) mobile phone applications were introduced in multiple countries over the course of 2020, for instance in the European Union [8]. These applications measure exposure to other individuals by using low-energy Bluetooth technology to identify their respective phones running the same or a compatible application. If tested positively, an index case can use the app to send notifications to potentially exposed individuals automatically who can then contact authorities, isolate themselves or get tested [9]. The major prospect of DigCT as compared to traditional CT is that infection chains might be broken sufficiently fast to contain an outbreak. In addition to the benefit of immediate notification upon case confirmation, and thus a reduced time until quarantine, DigCT may also identify contacts that are unknown to the index case, an advantage compared to manual contact tracing [10, 11]. While DigCT could have also had an indirect impact by providing access to anonymized, time resolved contact structures that would have made more informed mitigation strategies possible, a decentralized implementation was chosen in most countries due to privacy concerns, which makes this kind of data unavailable [12]. The success of such large-scale digital mitigation strategies mostly depends, for a voluntary and decentralized approach, on acceptance in the general population, proper usage of the application, and technical properties [13]. Numerous studies have already been conducted regarding benefits and limitations of DigCT concerning its use during the COVID-19 pandemic [14–27]. It was found that strong mitigation effects could only be observed with high app participation. From a contact network perspective, this is not surprising because DigCT is founded on sampling contacts in a population and, approximately, the fraction of sampled contacts scales with participation quadratically. So, for instance, if 10% of the population participates only 1% of all contacts occur between pairs of individuals that are users of a DigCT application. Nevertheless, no critical threshold exists below which DigCT had no impact on mitigation at all, suggesting that even a low proportion of app-participating individuals can cause a reduction in outbreak size [20]. In particular, DigCT might lead, in the absence of other NPIs, to an asymptotic exponential decay in prevalence, suggesting DigCT-induced “epidemic control” of an outbreak [11, 14, 23]. Yet, a quantitative assessment of the actual positive epidemiological effects and DigCT’s efficacy is challenging because a number of different variables are involved whose influence can be difficult to quantify empirically, e.g. participation rates, delays in quarantining and notification, the amount of traced contacts, high pre-/asymptomatic transmission rates, or missing bidirectional tracing [15, 18, 19, 23]. Recently, it was reported that a 30% participation rate might have lead to a 15% reduction in cases in the UK during the last quarter of 2020, while for France, a modeling analysis suggested an 8% reduction in peak prevalence (on top of household isolation, which was found to provide a base reduction of 27%) [22, 28]. For empirically recorded temporally resolved contact networks, a DigCT efficacy of ≲ 5% and ≈ 10% have been found based on modeling, respectively, for a participation rate of *a* = 30% [20]. It has been argued that further increasing the participation rate will lead to stronger mitigation or even containment [14, 22, 28]. The discrepancy of reported efficacies and the new availability of empirical data regarding how users interact with the respective applications warrant a deeper analysis of which factors influence the intervention’s success in which way.

In order to assess critical factors that determine the success of DigCT applications empirically, several field studies have been conducted, e.g. in Spain, [10], Norway [11], and Switzerland [29]. It was found that 64% (La Gomera, Spain) or 60.3% (Zurich, Switzerland) of all app users upload a positive test result. This is in agreement with the findings in Norway where 50%–70% of app users were reported to be active daily and may therefore be categorized as active users. In Germany, 62% of app users uploaded their positive test results up to May 20, 2021 [30]. Uploaded test results led to 6.3 (La Gomera) and 4.3 (Zurich) notified individuals per index case. Of those notified contacts, 10% and 53% reached out to authorities for follow-ups in La Gomera and Zurich, respectively. Both studies concluded a population-wide app-participation of around 30%. The proportion of notified contacts that are unknown to the index case ranges from 11% (Norway) to 20%–40% (La Gomera).

So far, it remains essentially elusive, however, to which extent DigCT applications may have contributed to the reduction of COVID-19 outbreak sizes. In particular, the empirical results discussed above have not been used to quantify outbreak reduction in real-world settings, apart from the UK [28]. Also, drastic non-pharmaceutical interventions (NPIs) usually referred to as “lockdown” measures most likely changed both the population’s contact structure as well as its mixing properties, hence the question arises how these changes, and contact structure in general, might influence the success of DigCT applications. Since DigCT relies on symptom-based testing to identify index cases, one may also wonder how its success depends on testing efficacy, which of both should be expanded to mitigate the disease’s spread further, and whether outbreaks can be prevented with higher levels of app participation.

To provide answers to these questions, we analyzed how strongly the introduction of DigCT via mobile phone applications can reduce the size of a COVID-19-like outbreak in different settings based on stochastic simulations on contact networks and considering the empirical results from real-world settings.

Our analysis suggests that in the absence of other NPIs, DigCT might lead to a decrease in outbreak size on the order of a few percent at best, which represents an upper bound as it ignores the fact that a large part of traced individuals are contacts that would have been found via manual contact tracing, too [10, 11]. DigCT might be more successful when contact structures are locally clustered, yet this advantage disappears if contacts per index case are broadly distributed, as is common for many social systems. We also find that the efficacy of DigCT increases with increasing symptom-based testing, but neither can prevent an outbreak of a disease for which 50% of infections are caused by pre- or asymptomatic individuals. We show that lockdown measures that reduce both contacts and mixing, paradoxically, make DigCT even less effective: supercriticality is crucial for DigCT to work. Consequently, containment and mitigation policies cannot rely on DigCT to prevent outbreaks, yet it might help complementing the manual CT process in times of rising case numbers.

## II. MATERIAL AND METHODS

We base our analysis on a stochastic dynamic disease model that accommodates the central mechanisms contributing to the outcome of DigCT applications. Contact structure is modeled by networks in which nodes represent individuals (*N* = 200,000) and links represent epidemiologically relevant contacts, see Fig. 1A. Given an app participation of 0 ≤ *a* ≤ 1, we sample ≈ *aN* individuals uniformly at random and mark them as app participants who can thus be traced. Susceptible individuals may be infected via contacts to infectious individuals, at which point they enter a latent phase and become presymptomatic infectious afterwards. In agreement with empirical results, we assume that 50% of infections are caused by individuals in the presymptomatic phase [4, 5, 31]. We include this to test the hypothesis that DigCT can reduce outbreaks by identifying and isolating presymptomatic cases quickly. Empirical studies have further shown that approximately 17% of infected individuals remain asymptomatic, while 83% become symptomatic eventually [32]. In both of these states, individuals infect non-isolated susceptibles, yet only symptomatic individuals can be identified via symptom-based testing, the major mechanism through which DigCT is invoked. Naturally, not every person with symptoms is tested for a variety of reasons. For instance, cases with mild symptoms may be less likely to seek medical help. Likewise, tests may be denied to certain individuals when maximum testing capacity is reached. When a symptomatic individual is tested, we assume that they are isolated and wait for their positive test result. When test results are received, app participants can choose to notify their contacts through the DigCT application which, according to the empirical studies cited above, ≈ 64% of app users do [10]. DigCT-participating contacts of such index cases are then notified and may choose to self-isolate or to get tested. We assume that 90% of contacts will self-isolate and only 10% will contact authorities to get tested themselves [10]. Susceptibles that have chosen to self-isolate return to their normal behavior after 10 days on average. Note that we distinguish between the number of documented, isolated infected individuals *X*(*t*), undocumented, non-isolated infected individuals *R*(*t*), and undocumented, self-isolated infected individuals *C*(*t*). The final size of an outbreak is therefore

**Figure 1.**
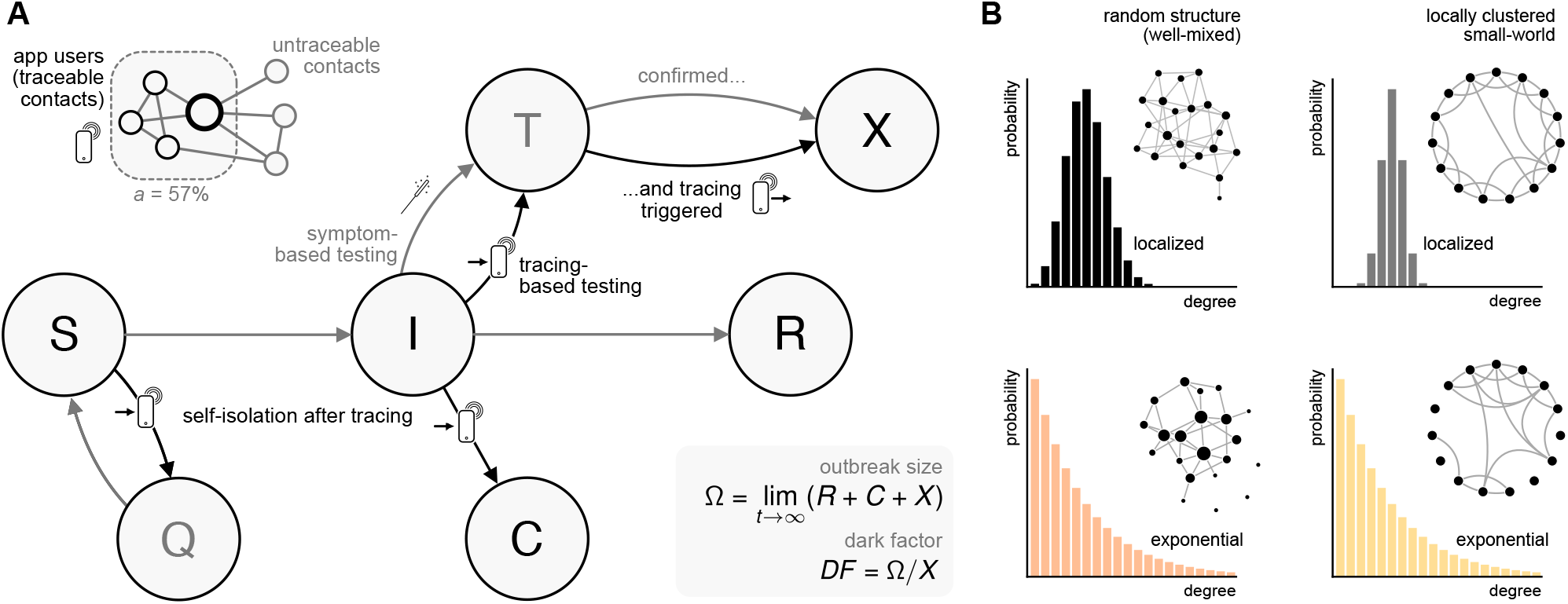
**A)** Symptom-based testing and DigCT. On a contact network, a proportion *a* of all individuals are marked as app users, chosen at random. Only contacts between app users are considered to be traceable. Susceptible individuals can be infected by presymptomatic, asymptomatic, or symptomatic individuals (*I*). Once infected, they enter a latent phase, where they are infected but not yet infectious (not shown here). Symptomatic individuals will be discovered either through symptom-based or app-induced testing, at which point they enter a latent testing state *T* that represents a waiting period during which the result is processed and prepared for upload. When a result is received, individuals enter the *X* state, at which point they may upload the result. Not all *T* individuals will upload their positive test result (either because they are not app users or because they decide not to). Those who do will trigger a notification of their contacts that are app users. We assume that notified app users will either self-isolate (enter the states *Q* or *C*) or get tested and may trigger contact-tracing themselves. In this case all, latent, symptomatic, pre-symptomatic, and asymptomatic contacts can be tested (in contrast to index-case testing, which is symptom-based only). Documented infections are counted in the *X* compartment, undocumented infections in the *R* and *C* compartments. Simulations are run until state-changing events no longer occur. The outbreak size and dark factor are given by Eqs. (1)-(2). **B)** We analyzed the efficacy of DigCT on four different network topologies to estimate the impact of different properties of real contact networks: (i) Erdős–Rényi networks (well-mixed and localized degree distribution), (ii) small-world networks (locally clustered with few long-range connections and narrow degree distribution) (iii) well-mixed networks with a broad degree distribution and (iv) locally clustered small-world networks with a broad degree distribution.

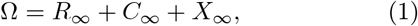

with *f*_∞_ = lim_*t*→∞_ *f* (*t*). We quantify the ratio of documented to all infections by means of the dark factor

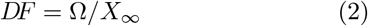

that is, if the documented number of infections is *X*_∞_, the actual number of infections is *DF* × *X*_∞_, i.e. *DF* times as large, also called the inverse ascertainment factor [28]. For constant symptom-based testing efficacy and varying app participation *a*, we refer to *DF*_0_ = Ω(*a* = 0)*/X*_∞_(*a* = 0) as the initial dark factor, i.e. the dark factor in the absence of DigCT.

Regarding the impact of contact network structure, we assume that the typical infectious period of the order of a few days is sufficiently long to model contact networks as an effective, averaged medium and simulate the dynamics on static networks only [33]. Due to data privacy reasons, it is impossible for researchers to record the complete contact structure between individuals using DigCT applications. We therefore test the efficacy of DigCT on multiple model network structures to assess how (i) broader degree distributions and (ii) strong local clustering affect the success of DigCT, both of which are typical properties of social networks [34]. We use four different network models to test DigCT efficacy on all combinations of the network properties (i) homogeneous and heterogeneous contact numbers and (ii) locally-clustered and well-mixed structure (see SI). Given the average contact number of 6.3 per index case and ≈ 33% app participation in the La Gomera experiment, we simulate networks with an average of *k*_0_ = 20 relevant contacts per person in the entire ensemble.

Additionally, we compared the efficacy of DigCT between two systems where (a) the disease can spread freely through a well-mixed contact structure and (b) “lockdown” measures reduce both the number of contacts as well as the population’s degree of mixing. Representing a “no-lockdown” scenario, we analyze an ensemble of Erdős–Rényi random networks with a mean contact number of *k*_0_ = 20 and a basic reproduction number of ℛ_0_ = 2.5. We assume that introducing lockdown measures reduces contacts by 50% and render clustering in contact structures more pronounced (a similar effect was observed in mobility networks [35]). Therefore, for a pure “lockdown” scenario, we simulate outbreaks on locally clustered small-world networks with *k*_0_ = 10 and ℛ_0_ = 1.25, implying a per-link transmission rate that is constant between both scenarios.

## III. RESULTS

Assuming an app participation of *a* = 30% and an initial dark factor of *DF*_0_ = 4 (which implies a 30% detection rate of symptomatic individuals [36, 37], see SI), we find that DigCT alone leads to reductions in outbreak sizes on the order of ≈ 5%. This result is robust for homogeneous well-mixed networks as well as networks with broader contact distributions. Only locally clustered networks with narrow contact distributions yield a higher reduction of ≈ 12% (see Fig. 2A). However, this increased reduction vanishes when broader contact distributions are considered in the presence of local clustering. Increasing the efficacy of symptom-based testing reduces the dark factor and enhances the relative success of DigCT, which remains on a very low level nevertheless (Fig. 2A). The dark factor itself changes only marginally when increasing app participation (see SI Fig. 6B), implying that DigCT will not contribute towards a clearer picture of the true extent of an outbreak. The epidemic threshold is unaffected by DigCT and its positive effect does not increase substantially for other values of ℛ_0_ (see SI Fig. 6A).

**Figure 2.**
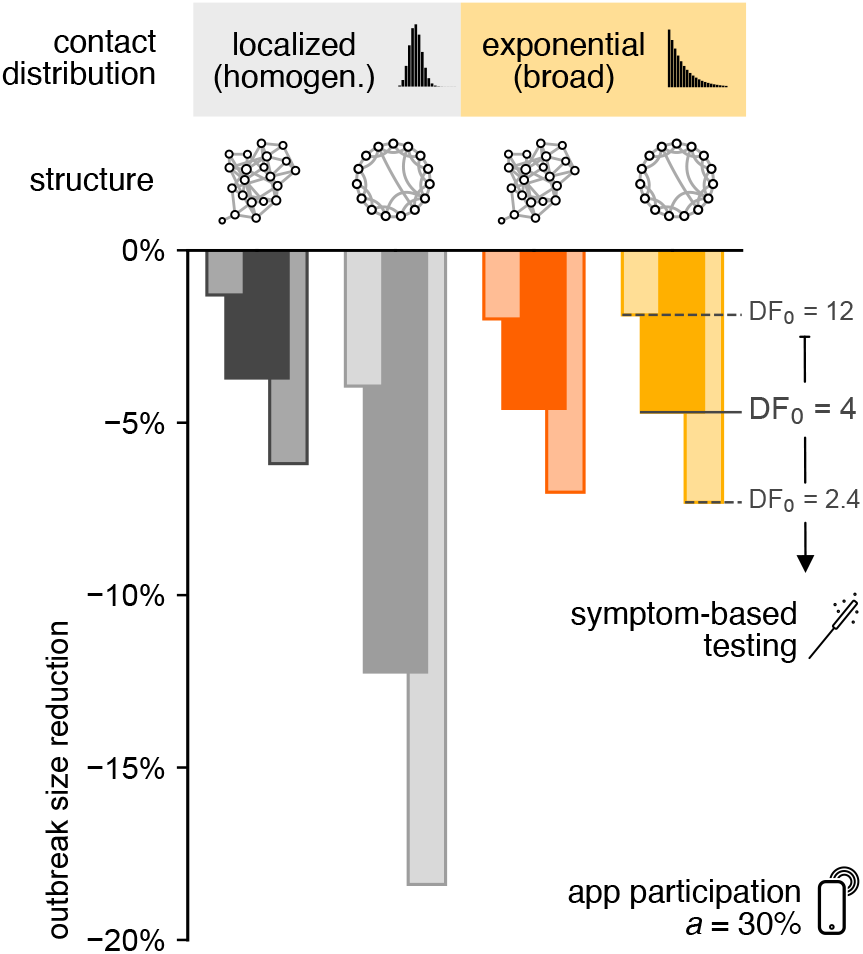
Relative mean outbreak size reduction 1 − ⟨ Ω(*a* = 30%) ⟩ */⟨* Ω(*a* = 0) ⟩ caused by DigCT. The app participation rate was fixed at *a* = 30%, and symptom-based testing was assumed to lead to initial dark factors of *DF*_0_ ∈ {12, 4, 2.4} (note that ⟨Ω(*a* = 0) ⟩ depends on the initial dark factor as well as the contact structure, see Fig. 4). DigCT has a stronger effect in locally clustered networks with narrow degree distributions as compared to their random counterparts (see grey bars). To test whether this success prevails for systems that resemble real social networks more closely, we further measured outbreak size reduction for networks with heterogeneous contact distribution. We find that the advantage that local clustering has over random structures vanished when one additionally considers broader contact distributions (see orange bars). Increasing symptom-based testing (i.e. decreasing the dark factor) enhances the efficacy of DigCT. All network structures were simulated with an average contact number of *k*_0_ = 20 per individual.

**Figure 3.**
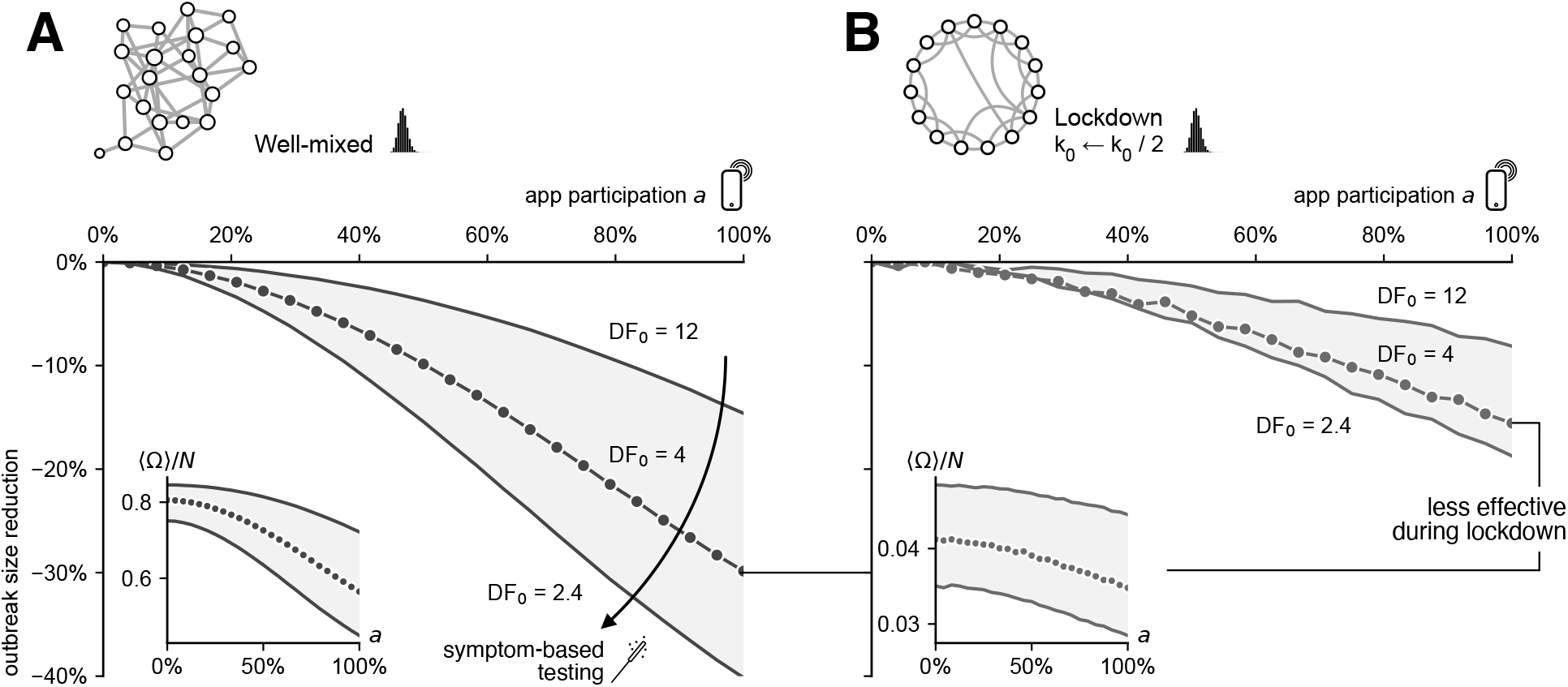
Capturing “lockdown” measures in our model, we compare the relative mean outbreak size reduction in a situation where **(A)** the disease spreads freely to a situation where **(B)** the disease’s spread is mitigated by (i) reducing the number of contacts by 50% and (ii) assuming a locally more clustered contact structure. Despite what our result of Fig. 2 implied, the locally clustered structure does not enhance DigCT’s success when the disease is subcritical. This suggests that supercriticality is necessary for DigCT to work more efficiently.

Considering lockdown measures that reduce both the number of contacts as well as mixing in the population, we compared the DigCT-induced relative outbreak size reduction for two different networks, one where the disease can spread freely through a well-mixed contact structure and one where a “lockdown” halved the number of contacts and exposes local clustering, effectively turning the infectious disease dynamics subcritical. We observe a much lower relative reduction in the outbreak size in the lockdown scenario than in the free scenario, even though contacts have been reduced and our previous analysis suggested increased clustering or decreased mixing may enhance DigCT’s success (Fig. 2B). This analysis demonstrates that DigCT efficacy is directly related to the number of infectious contacts and hence prevents more infections in supercritical situations (both absolutely and relatively).

Many countries experienced similar, more complex patterns of pandemic trajectories up until now: Often, periods of un- or partially mitigated spread were followed by suppressed growth, stifled by “lockdown” measures that temporarily reduced the effective reproduction rate. To investigate how the dynamic trajectory of an epidemic influences the number of infections averted by DigCT, we compared epidemics that (i) spread freely and (ii) were dominated by periodic changes of unmitigated growth and suppression by other NPIs (see SI). We find that the success of DigCT is strongly influenced by the dynamics of the pandemic (see SI, Fig. 11). As a rule of thumb, DigCT will prevent more cases in phases of epidemic growth. Yet, after an outbreak reaches its peak and if no other NPIs mitigate the spread, prevalence will decay less quickly in DigCT-mitigated systems than if herd immunity was reached naturally, which, in turn, leads to a negative number of cases averted during this phase of the outbreak, reducing the overall percentage of averted infections until the epidemic is over. This reduction in efficacy can be avoided when other NPIs strongly suppress further dissemination of the disease, in which case the average number of averted cases can, but does not have to, remain at higher values. This effect is more pronounced in systems where testing is not symptom-based (see SI, Fig. 12).

To analyze whether increasing app participation or increasing efficacy of symptom-based testing has a higher impact on mitigation we compared how much the outbreak size is reduced if either the dark factor is reduced from *DF*_0_ = 4 to *DF*_0_ = 2.4 (by detecting 20% more symptomatic individuals, see SI) or the app participation is increased by 20% (from 30% to 50%). In well-mixed networks and networks with an exponential degree distribution, neither increase has an advantage over the other (Fig. 4). In locally clustered contact structures with homogeneous contact numbers, increasing symptom-based testing reduces the outbreak size more strongly. The benefit of increased symptom-based testing in locally clustered structures can thus only be maintained if the number of contacts is homogeneous, which is untypical for social networks. Neither increase will lead to containment.

**Figure 4.**
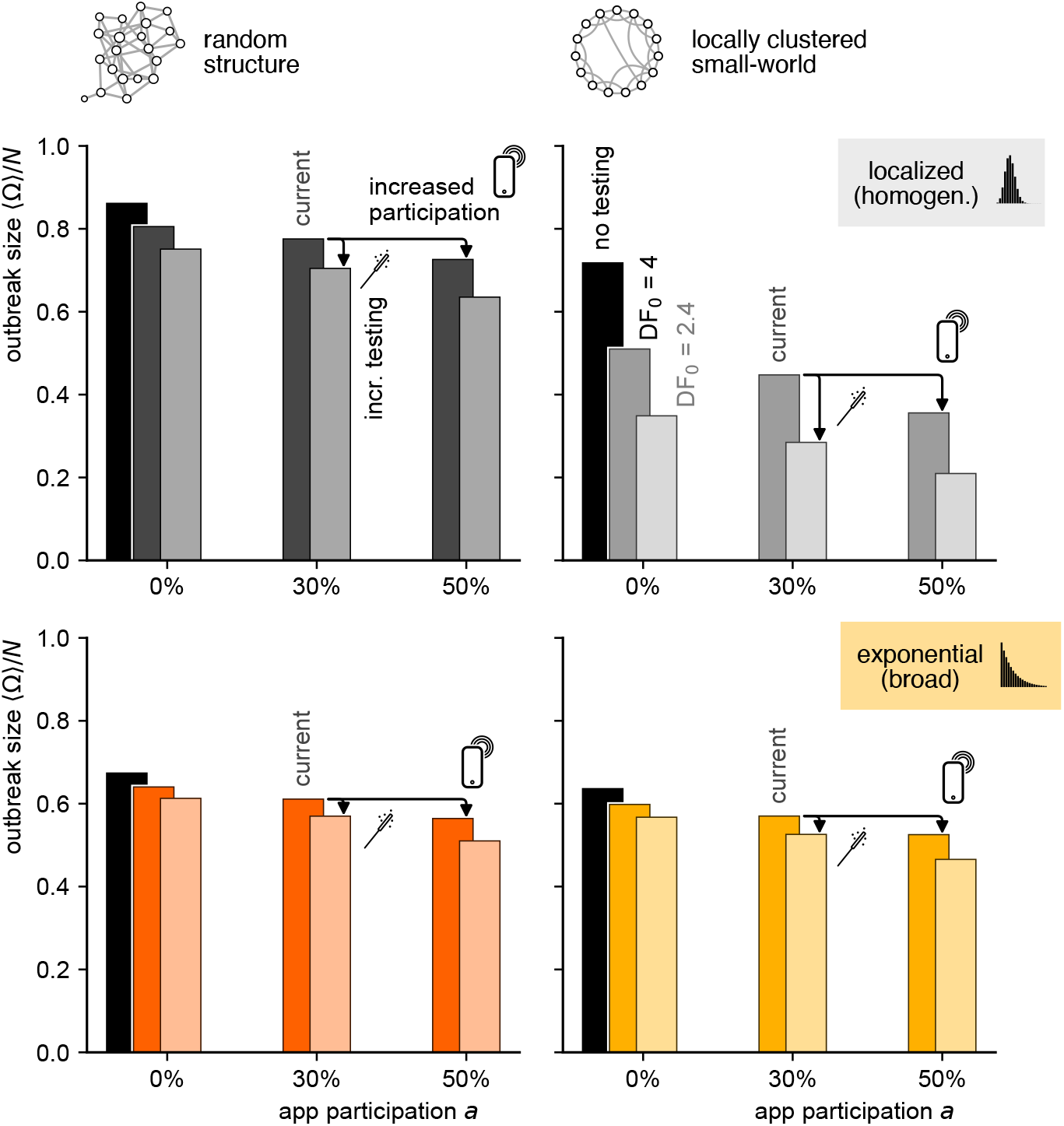
Comparison of the absolute outbreak size ⟨ Ω ⟩*/N* for different network models introduced in Fig. 1, shown for app participation values of *a* ∈ {0%, 30%, 50%}. We further compared the absence of symptom-based testing, and testing that would lead to dark factors of *DF*_0_ = 4 and *DF*_0_ = 2.4, respectively. Empirical observations suggest that several countries are currently in a state of an *a* ≈ 30% app participation rate and a dark factor on the order of *DF*_0_ = 4. We find that for three of the four network structures no difference in increasing either symptom-based testing or app-participation. For locally clustered small-world networks with narrow contact distribution, an increase of symptom-based testing leads to a stronger reduction than an increase in DigCT participation.

Since we consider that susceptible contacts will isolate themselves for an average of 10 days after receiving a notification, one may wonder how strongly DigCT-induced mitigation depends on a considerable number of susceptible individuals shielding themselves from the infection process. Indeed, considering that susceptibles would not isolate themselves, we find a reduced efficacy in outbreak reduction (yet barely effecting our results). Similarly, the mitigation effect is reduced if less contacts would respond to a notification (see SI Fig. 7).

So far, we assumed that only 10% of traced contacts get tested and hence may induce next-generation tracing, as based on the empirical results found in Spain. This number was measured, however, in a situation where the population was disease-free. In a real outbreak situation, one might assume that this value increases due to an increase in perceived risk. Considering that 50% of all traced contacts initiate further tracing [29], DigCT efficacy does not increase substantially for dark factors on the order of *DF*_0_ = 4 or app participation on the order of *a* ≈ 30%. However, for high dark factors (i.e. low symptom-based testing efficacy) or high app participation, an increased probability to induce next-generation tracing increases DigCT’s relative efficacy more strongly (see SI Fig. 8).

## IV. DISCUSSION

Contrary to the positive expectations DigCT has raised initially, we conclude that its impact on the containment of COVID-19 outbreaks was low in most countries. While our results suggest a relative reduction of case numbers in the single-digit percentages for otherwise unmitigated outbreaks, we ignored the fact that many of the cases that are found via DigCT will be found via manual CT, as well (by authorities or by self-induced household isolation), as suggested by the empirical studies in Spain and Norway [10, 11]. The initial claim that the introduction of DigCT might lead to “epidemic control” [14] is thwarted by the fact that the intervention has a higher efficacy in phases of strong growth and higher prevalence.

Our analysis demonstrates, however, that DigCT does indeed prevent cases, even if this number is in the low percentages. Since every prevented case is a life potentially spared, one may argue that its implementation is of use in any case. Yet, the consequences of the intervention’s introduction on the behavior of the population should not be disregarded. Since DigCT applications have been marketed to be rather effective, their introduction may have lead to an increase in contacts, which in turn could nullify the method’s benefits.

The efficacy can be enhanced by increasing participation rates, randomized or symptom-based testing, the proportion of test results uploaded to the app, the proportion of contacted people that trigger next-generation tracing, as well as the introduction of strict NPIs that strongly suppress growth after an outbreak emerged. It is questionable, however, whether higher participation rates than those observed can be achieved because the number of people eligible for participation in DigCT is limited regardless [38].

In our analysis, we disregarded that contact structures and app participation may assume heterogeneous values across different regions and age groups. We expect that local clustering of app usage will improve the method’s efficacy due to Jensen’s inequality. In the UK, higher values of relative outbreak size reduction have been reported [28], which might be attributed to technical differences of the application, the authors ignoring the influence of the testing-dependent dark factor, or that the impact of DigCT was measured exclusively over a period where cases mostly grew, which is when the relative effect of DigCT will be stronger.

Apart from their low impact on containment, DigCT applications might have been an outstanding possibility for researchers in digital epidemiology to learn more about temporal and spatial contact structures by allowing access to aggregated, anonymized contact histories. Doing so would have enabled the field to give more targeted statements about the contact structure-mediated risk of infection. Unfortunately, this has not been made possible because due to privacy concerns, a decentralized approach was chosen in most countries, in which such data is unavailable. Given our analysis, one might argue that this kind of information, that comes as a by-product of DigCT, might have had a similar impact on containment than the applications’ initial purpose, even if less privacy would have implied lower participation rates.

In summary, despite the promising outlook DigCT applications had initially, we argue that at best they can support manual contact tracing when outbreaks become large, but will not mitigate outbreaks significantly, which is why policy makers should concentrate on other non-pharmaceutical interventions for the containment of COVID-19 or similar diseases.

## Data Availability

No empirical data has been collected in this study.

## ACKNOWLEDGMENTS

BFM is financially supported as an *Add-On Fellow for Interdisciplinary Life Science* by the Joachim Herz Stiftung. BFM thanks F. Klimm and F. Schlosser for helpful discussions. DB would like to thank I. Mortimer for valuable comments on the manuscript. AB thanks G. Saf for valuable discussions.

## Appendix A: Model

We devise a stochastic infection model based on the compartmental susceptible-exposed-infectious-removed (*SEIR*) model [2]. We split the infectious state to account for presymptomatic (*I*_*P*_), asymptomatic (*I*_*A*_) and symptomatic (*I*_*S*_) infectious individuals in order to adequately test the hypothesis that DigCT can contribute to containment by quickly identifying pre- and asymptomatic individuals. Furthermore, we introduce states for individuals being susceptible and quarantined (*Q*), infected and tested (*T*), known infected and quarantined (*X*), and unknown infected and quarantined (*C*), see Fig. 5. The model is run on static networks of node set size *N* and mean degree *k*_0_. The following events may happen.

**Figure 5.**
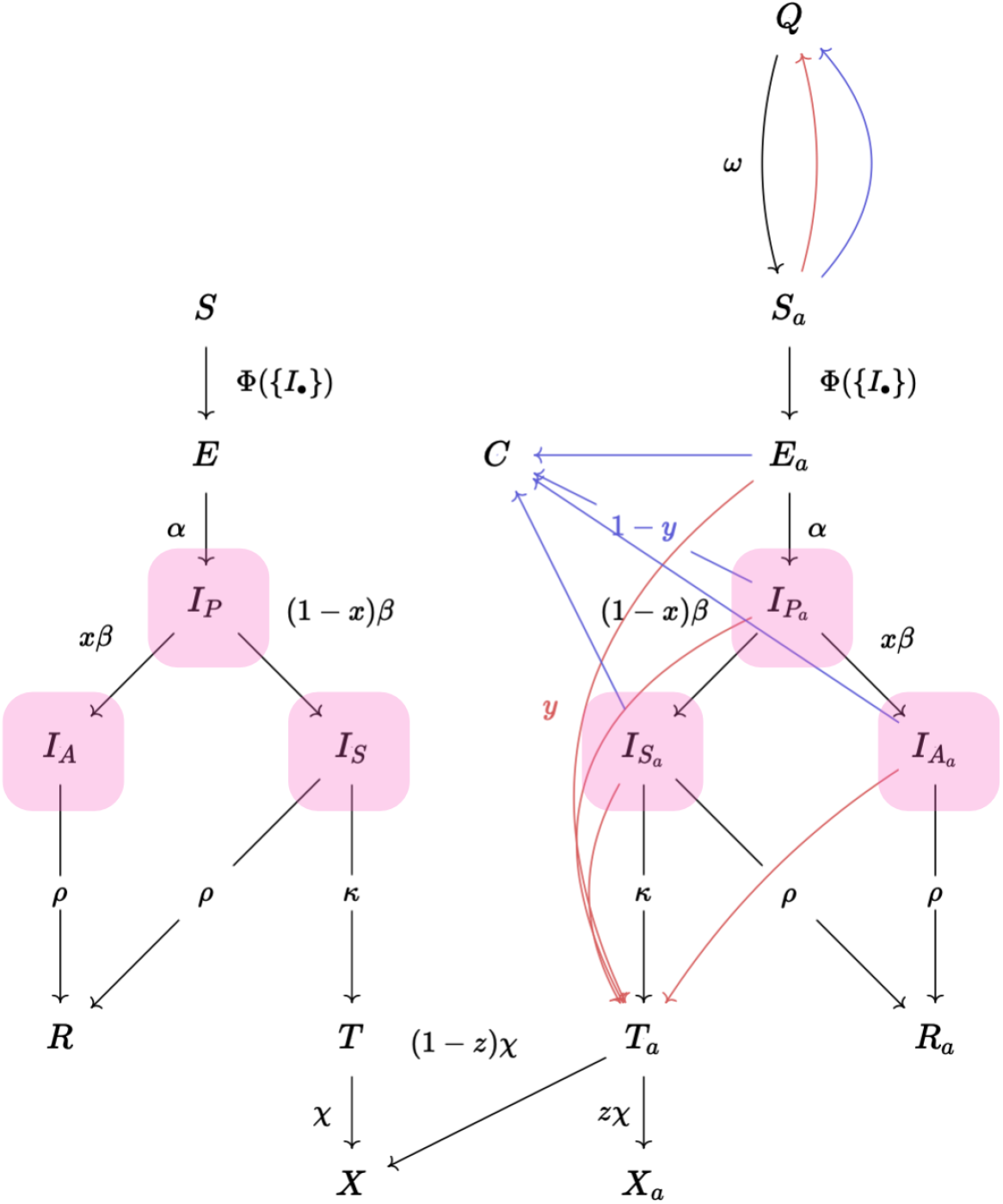
Flowchart of the epidemic digital contact tracing model used in this study. States of app-participating individuals are marked with subscript *a*. Susceptible individuals *S* (*S*_*a*_, respectively) will become exposed *E* (*E*_*a*_) after getting infected and enter a presymptomatic infectious state 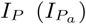. Susceptible individuals can be infected by any neighbor in any infectious state (*I*_*A*_, *I*_*P*_, *I*_*S*_, *I*_*A,a*_, *I*_*P,a*_, *I*_*S,a*_). After being presymptomatic infectious, individuals become either asymptomatic 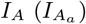 or symptomatic infectious 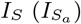 and can either recover *R* (*R*_*a*_) or, if they are symptomatic, they can be found by symptom-based testing *T* (*T*_*a*_). Tested individuals that are not app participants (*T*) will enter the final infected and quarantined compartment *X*. Tested individuals that are app-participants *T*_*a*_ will either upload their positive test result or not. If they do, they enter the *X*_*a*_ state, if not they behave like non-participating individuals and enter *X*. When an individual enters *X*_*a*_, its infected neighbors will either choose to self-quarantine *C* or to get tested *T*_*a*_ to induce next-generation tracing. Susceptible neighbors will always choose to self-quarantine (*Q*) and will re-enter society after time *ω*^*−*1^.

- 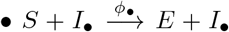: Susceptible individuals will become exposed after getting infected by any of their a-, pre- or symptomatic neighbors. A substantial proportion of infections are caused by pre- and asymptomatic transmissions [4, 5, 31]. We assume that transmissions caused by presymptomatic individuals represent half of all infections occured and that asymptomatic and symptomatic individuals are equally infectious (while asymptomatic infections may be associated with lower viral shedding, it has been reported that they might be equally infectious nevertheless [39], furthermore, asymptomatic infecteds are less likely to change their behavior, in contrast to symptomatic individuals who will be more prone to self-isolation). In the base parameter set, a single infectious individual in an otherwise susceptible population transmits the disease to ℛ_0_ = 2.5 susceptibles. Of these transmissions, ℛ_0_*/*2 happen during the presymptomatic phase and ℛ_0_*/*2 in either an asymptomatic or a symptomatic phase. To make simulations comparable across network models, we gauge the transmission rate per link using the mean-field definition of ℛ_0_ as

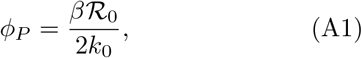

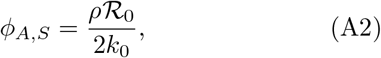

where *β*^*−*1^ is the average duration of the presymptomatic infectious period and *ρ*^*−*1^ is the average duration of the remaining infectious period (see below for numerical values).
- 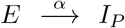: Exposed individuals will become presymptomatic and therefore infectious after a latency period *τ*_*E*_ = *α*^*−*1^ = 3 d [31, 40–44].
- 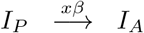: A proportion *x* = 0.17 [32] of all presymptomatic individuals will not develop symptoms and will therefore become asymptomatic infectious after an average time *τ*_*P*_ = *β*^*−*1^ = 2 d [31, 42].
- 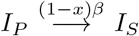:A fraction (1 − *x*) of all presymptomatic individuals will develop symptoms after an average duration of *τ*_*P*_ and will therefore be symptomatic infectious.
- 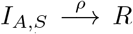 : After an infectious period of *τ* _*I*_ = *ρ*^*−*1^ = 7 d [45, 46] asymptomatic and symptomatic individuals will be removed (*R*) from the dynamics.
- 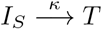 : With symptom-based testing, a symptomatic infectious individual can be detected with rate *κ* which is determined by the isolation probability *q* = *κ/*(*κ* + *ρ*), which measures the probability to detect a symptomatic individual before it recovers. Hence, *q* quantifies the efficacy of symptom-based testing: For higher values of *q*, more symptomatic individuals will be detected earlier. The detected individual will enter the state *T*, which acts as a waiting compartment and symbolizes the time that passes between detection, receiving, and uploading a positive test result to the app if the individual is an app participant.
- 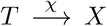 : After being detected, the individual will enter the final infected and quarantined state *X* after a time *τ*_*T*_ = *χ*^*−*1^ = 2.5 d. We base this assumption on the following considerations: If on day 1 a sample was taken, then on day 2 the test result will be available and can be uploaded. Then on day 3 the contacts of the confirmed case will download the new data and receive a notification. When entering *X*, the app-participating individuals can induce DigCT (red and blue arrows in Fig. 5).
- DigCT: Digital contact tracing is modeled as a conditional stochastic event that happens immediately after a *T*_*a*_ → *X*_*a*_ event is triggered. The probability of an app-participating confirmed infectious individual to upload a positive test result when entering *X*_*a*_ is assumed to be *z* = 0.64 [10, 29]. All of this individual’s neighbors will be notified and will be quarantined. Yet, only a fraction *y* = 0.1 will induce further tracing themselves (if they are infected). This implies that if an individual in *T*_*a*_ enters the *X*_*a*_ state and uploads its result, its infected (any of *E*_*a*_, *I*_*P,a*_, *I*_*A,a*_, *I*_*S,a*_) notified neighbors are entering the *T*_*a*_ state with probability *y* (red in Fig. 5), and are therefore counted as known to the public health system. Otherwise, they enter the *C* state with probability 1 − *y* (blue in Fig. 5). Susceptible notified neighbors will be quarantined (entering *Q*) in a manner similar to infected neighbors, but will never induce further tracing and will return to their normal behavior (*S*) after an average duration of *τ*_*Q*_ = *ω*^*−*1^ = 10 d [47]. The remaining 1 − *z* confirmed infectious app-users that will not upload their test result will behave as none-participating individuals and enter *X*. Simulations with only half of the individual’s contacts reacting, or *y* = 0.5, or changing the base configuration in a way that susceptible contacts will never be isolated can be found in App. C.

We use an adapted version of Gillespie’s algorithm to simulate the link- and node-mediated processes listed above, which is a generalized version of the rejection-sampling algorithm presented in [48]. The model and simulations were implemented using the infectious disease modeling framework “epipack” [49]. All simulations were initiated with *I*_*P*,0_ presymptomatic infectious individuals, where *I*_*P*,0_ was drawn from a binomial distribution with size *N* and mean *N/*100. For each simulation, a number of app participants was drawn from a binomial distribution with size *N* and mean *aN*. Both initially infected and app participants are drawn uniform at random from the node set. For each parameter set and structure 100 independent simulations were run until the total event rate reached zero, marking time *t*_f_. Subsequently, the final outbreak sizes were computed as *X*_∞_ = *X*(*t*_f_) + *X*_*a*_(*t*_f_), *C*_∞_ = *C*(*t*_f_) + *C*_*a*_(*t*_f_), and *R*_∞_ = *R*(*t*_f_) + *R*_*a*_(*t*_f_) for each simulation (note that *f* (*t*) symbolizes the number of individuals that are in state *f* at time *t*).

## Appendix B: Network Models

Simulations were performed on four different static network models representing several properties of empirical social contact networks. For each simulation, a network instance was sampled from each of the respective network model ensembles, on which the simulation was then run. We used *N* = 200 000 nodes and an average number of contacts (average degree) of *k*_0_ = 20 if not stated otherwise. While the results converge for lower values of *N*, this value was chosen to strongly reduce the uncertainty of displayed averages. As a base model we sample Erdős–Rényi/Gilbert *G*(*N, p* = *k*_0_*/*(*N* − 1)) random networks [50–52]. These networks represent well-mixed systems with homogeneous contact distributions. For locally clustered small-world networks with homogeneous contact distribution, we constructed Watts–Strogatz-like small-world networks with an algorithm that draws edges according to a distance-dependent connection probability with a short-range kernel [53, 54] (here, distance refers to lattice distance on a one-dimensional ring). We chose a long-range probability redistribution parameter of *β* = 10^*−*7^ where the random walk mixing time is on the order of that of random networks but average clustering is high (𝒞 = 0.7). In order to represent well-mixed systems with heterogeneous contact distributions, we generated degree sequences following an exponential distribution and constructed networks using the configuration model [55], where we removed self-loops and duplicate links. Exponentially distributed contact numbers have been observed in several human contact networks [33, 56]. While systems with heterogeneous contacts are often modeled with power-law degree distribution, an exponential distribution suffices to induce typical spreading-related properties such as a lowered epidemic threshold and lower endemic states for higher infection rates [33]. An exponential contact distribution also ensures that the maximum number of contacts in a network of *N* = 2 × 10^5^ nodes is on the order of Dunbar’s number [57].

Last but not least, we want to represent systems that are locally clustered, small-world, and have a heterogeneous contact distribution. We developed a new model that meets these properties. Networks are constructed as follows. Arrange *N* nodes on a ring. Each node is assigned a number of stubs from an exponential distribution. Now iterate over all nodes in one of three orders: (i) by descending stub count, (ii) by ascending stub count, or (iii) randomly. For each node *u*, iterate over all nodes *v ≠ u* on the ring, sorted by lattice distance to the focal node *u*, starting with the nearest node. If node *v* has stubs left and *u* is not yet connected to *v*, connect to node *v* and move on to the next node. Iterate until *u* has no stubs left or until the node that is farthest away (lattice distance *N/*2) has been visited. If focal nodes *u* are iterated in descending order, hubs are connected first and will likely find locally available stubs that belong to nodes of small stub count. At last, a final number of nodes with small stub counts will have to connect to other low-degree nodes that are far away. Therefore, the structure is dominated by local connections and a small number of low-degree nodes will have non-local connections. The generated network will resemble a lattice, but nodes will have exponentially distributed degrees. Since high-degree nodes are connected first and to nodes with low stub count, degree assortativity will be negative. The probability that an edge connects two nodes at lattice distance *d* will be concentrated at low values of *d*. An empirical analysis reveals that a small amount of edges will connect far-away regions, yet *d* will not reach values of maximum distance *N/*2 (see Fig. 9).

If focal nodes *u* are iterated in ascending order, low-degree nodes are connected first and “fill up” the local connections such that once high-degree nodes are connected, only long-range connections are possible, and with high probability only to other high-degree nodes. Therefore, hubs will play a mixing role, connecting different regions of the network, while low-degree nodes will contribute a lattice-like, highly clustered structure. Degree assortativity will be positive. The probability that an edge connects two nodes at lattice distance ≤ *d* approximately follows *d*^*−*1^ (see Fig. 9).

If focal nodes *u* are iterated in random order, low-degree nodes are connected first with higher probability (because there are significantly more low-degree nodes). However, low-degree nodes will also connect last with higher probability. Hence, nodes of any degree will form a lattice-like structure, while nodes of any degree will play a mixing role, connecting far-away regions. Degree assortativity will be close to zero. The probability that an edge connects two nodes at lattice distance ≤ *d* approximately follows *d*^*−*1^ (see Fig. 9).

Since social networks tend to have positive degree assortativity [58, 59], we choose to construct networks in the “ascending” order only. In Fig. 10, we compare a single result on these networks to networks that were created using the “random” order to find that degree assortativity makes no substantial difference.

We relied on networkx [60], numpy [61], and matplotlib [62] for additional analyses and illustrations.

## Appendix C: Additional Analyses

### 1. Varying basic reproduction number

To make sure that our analysis does not rely on the chosen basic reproductive number and to study whether DigCT influences the epidemic threshold of the system we compared the mean and coefficient of variation (CV) of outbreak size for ℛ_0_ ∈ [0.1, 10] for *a* = 0 (no DigCT) and *a* = 30% app participation (see Fig. 6). For both, (i) a random network with exponentially distributed degree as well as (ii) a homogeneous, locally clustered network, minor differences in outbreak size and no differences in CV were observed for the whole range of ℛ_0_ suggesting that the efficacy does not increase substantially for other ℛ_0_ and that the epidemic threshold is not changed, respectively.

**Figure 6.**
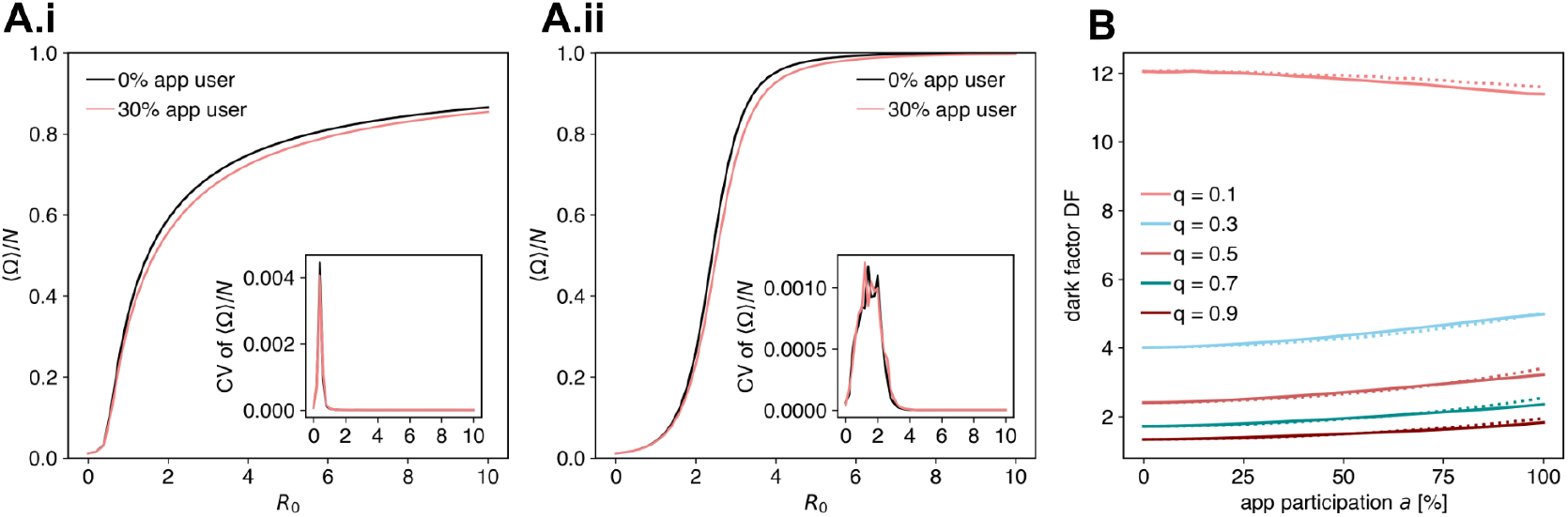
Mean and coefficient of variation (CV) of outbreak size ⟨ Ω ⟩*/N* for increasing reproduction number ℛ_0_ ∈ [0.1, 10] with *DF*_0_ = 4 and *a* = 0 (no DigCT) or *a* = 30% app participation in the population on **(A.i)** a random network with an exponential degree distribution and **(A.ii)** a locally clustered small-world network with a localized degree distribution. **(B)** Dark factors caused by increasing efficacies of symptom-based testing on (solid line) a random network with an exponential degree distribution and (dotted line) a locally clustered small-world network with a localized degree distribution. For *q* ≥ 0.3 the dark factor increases with rising app participation. This is due to the increasing number of individuals that are being traced but choose to self-quarantine and therefore remain undocumented.

Increasing the efficacy of symptom-based testing decreases the dark factor but with increasing app participation the dark factor shows only minor changes for both networks (see Fig. 6B). This suggests that increasing DigCT does not contribute to obtaining a clearer picture of the outbreak. We attribute this finding to the fact that many contacts choose to self-quarantine instead of getting tested as described above.

### 2. Influence of proportion of contacts that react to notification

The assumption that all notified contacts react reflects an upper bound. Lower participation should be considered. Hence, we have simulated that instead of all, only 50% of all contacts react after a notification. This means of all infected notified contacts 5% (instead of 10%) will get tested and 45% (instead of 90%) will choose to self-quarantine, while the remaining contacts will not change their state. Of all susceptible notified contacts 50% (instead of 100%) will choose to self quarantine (see Fig. 7ii). We find that doing so leads to smaller values of relative outbreak size reduction, especially for higher dark factors (i.e. less efficient symptom-based testing).

**Figure 7.**
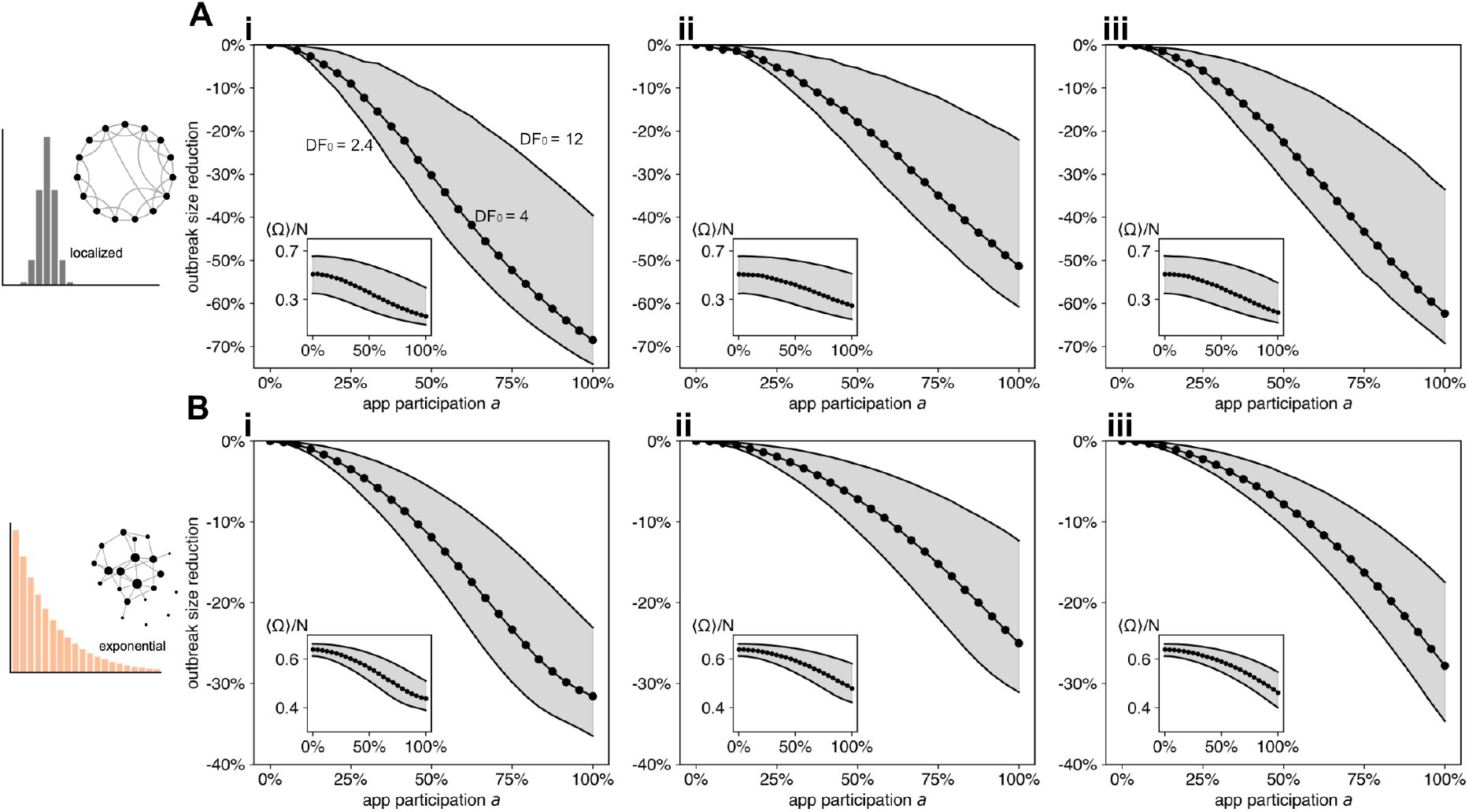
Outbreak size ⟨ Ω ⟩*/N* and relative outbreak size reduction caused by DigCT with *DF*_0_ ∈ {12, 4, 2.4} for increasing app participation *a* for **(i)** the base parameter set, **(ii)** with only 50% of traced contacts reacting to a notification and **(iii)** without isolation of susceptible contacts in **(A)** a locally clustered small-world network with a localized degree distribution and **(B)** a random network with an exponential degree distribution.

### 3. Estimating the contribution of isolation of susceptibles

Additionally, one might wonder how reducing the pool of susceptibles by isolating them contributes to the efficacy of DigCT. Hence, we have also simulated that none of the susceptible contacts choose to self-quarantine. Doing so reduces the efficacy of DigCT, but not dramatically. The change is slightly more pronounced for mid-range values of the app participation *a* (see Fig. 7iii).

### 4. Influence of next-generation tracing

In our base analysis we have chosen that 10% of notified infected contacts will get tested and could therefore induce further tracing as it was found on La Gomera [10]. We have also discussed that higher values were found in Zurich [29]. We therefore reran simulations with *y* = 0.5, i.e. 50% of notified contacts will get tested, to analyze the impact of this parameter. We simulated on well-mixed and locally clustered networks (both with exponential degree distribution) how results are changed if (instead of 10%) 50% will get tested (see Fig. 8). Increasing the proportion of traced individuals that get themselves tested to 50% is most beneficial for high dark factors while with low dark factors only minor differences were observed (tested on random networks). These differences are also visible in the small-world network but are not as pronounced.

**Figure 8.**
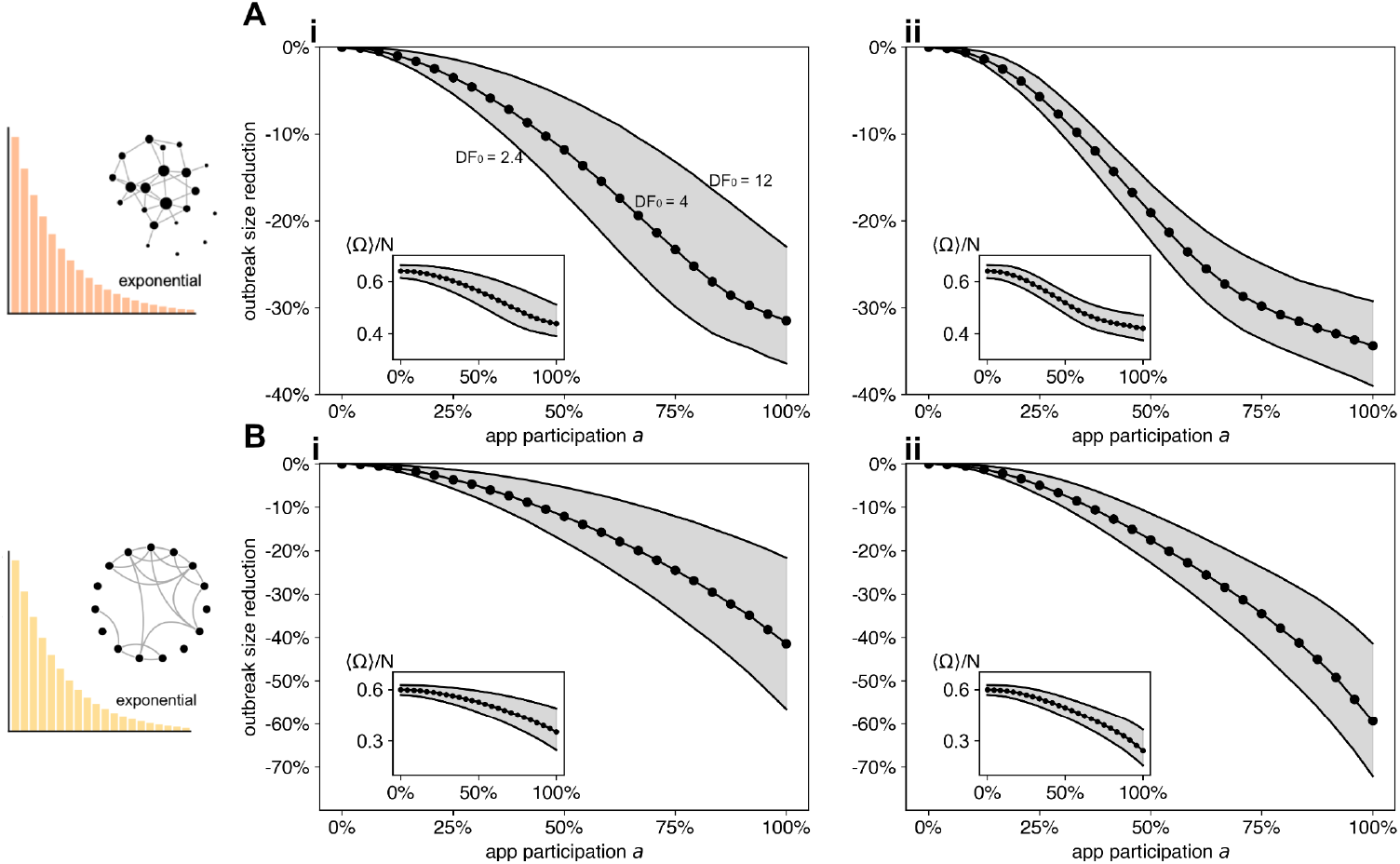
Outbreak size ⟨ Ω ⟩*/N* and relative outbreak size reduction caused by DigCT with *DF*_0_ ∈ {12, 4, 2.4} for increasing app participation *a* in **(A)** a well-mixed structure with an exponential degree distribution and **(B)** in a locally clustered small-world network with an exponential degree distribution where **(i)** 10% (*y* = 0.1) and **(ii)** 50% (*y* = 0.5) of traced infected contacts can induce further tracing.

**Figure 9.**
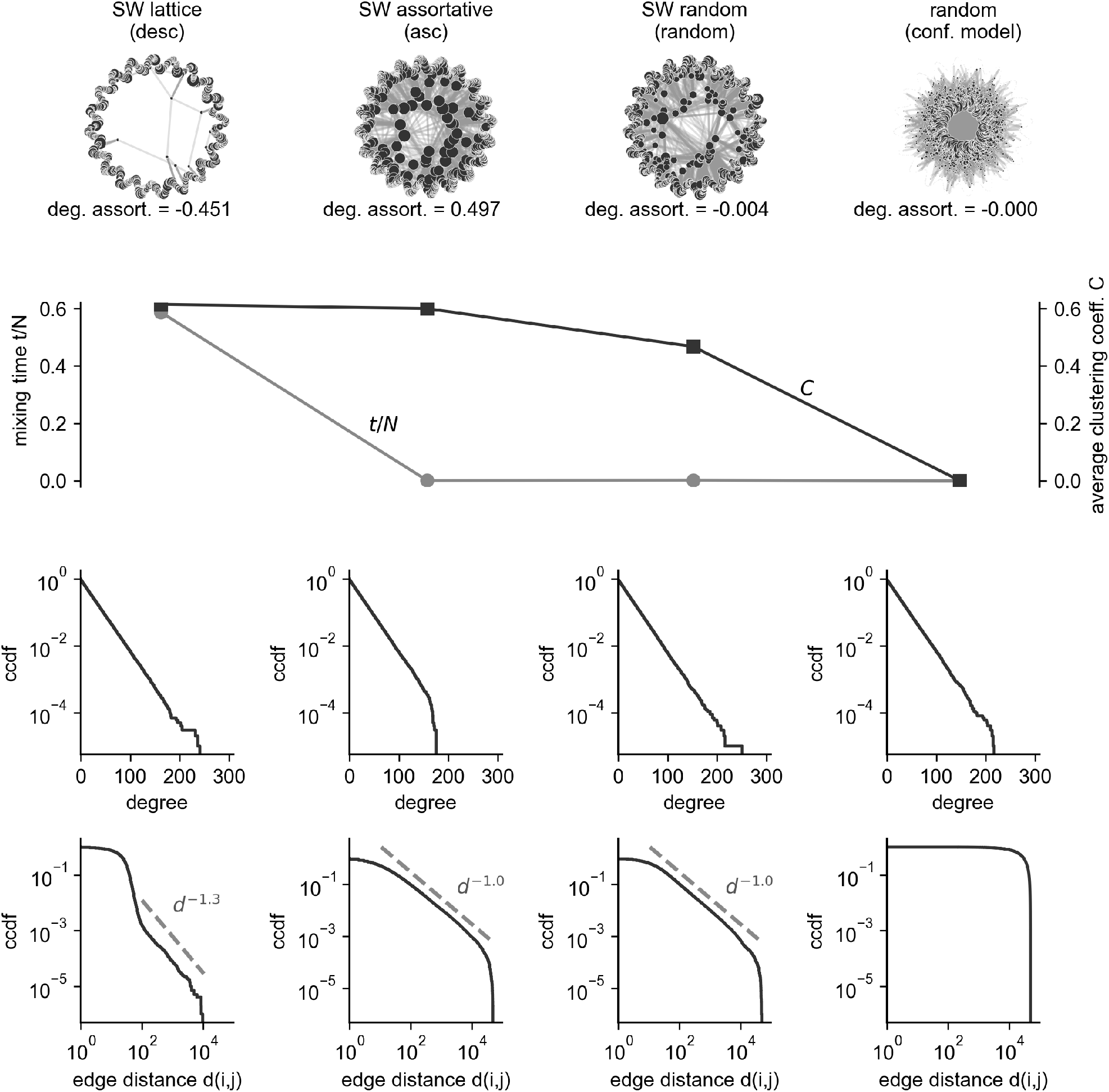
Illustration of network properties of the small-world model with heterogeneous degree distribution described in App. B. Networks can be created by iterating over nodes in “descending”, “ascending” or “random” order. We compare the structures to a well-mixed model with similar degree sequence (configuration model). The upper row shows illustrations of the networks that are constructed using the proposed algorithm. Larger disks represent nodes with higher degree and nodes are positioned closer to the center proportional to the maximum distance that any of its connecting edges bridges. For these illustrations, networks with *N* = 5000 and *k*_0_ = 10 were generated. For the rows below, a single network instance for each model was analyzed, with *N* = 100,000 and *k*_0_ = 10. We use the per-node random walk mixing time of the network’s largest connected component [53] and the average local clustering coefficient 𝒞 [63] to illustrate the small-world effect. The value of 𝒞 remains high for all models that are constructed using this algorithm. When iterating nodes in “descending” order, the mixing time is significantly larger than iterating in “ascending” or “random” order, both of which yield mixing times on the order of the random network (configuration model). For all network models, the degree distribution is approximately equal (note that for the “ascending model”, however, the maximum degree is not as high as for the other models). For the “descending” model, edges mostly bridge short distances, with a few edges connecting regions that are further away (yet not reaching the maximum distance *N/*2). The remaining two models yield edge distances *d* where the complementary cumulative distribution function (ccdf) follows a power-law *d*^*−*1^. In contrast, the existence of an edge does not depend on distance in the configuration model.

**Figure 10.**
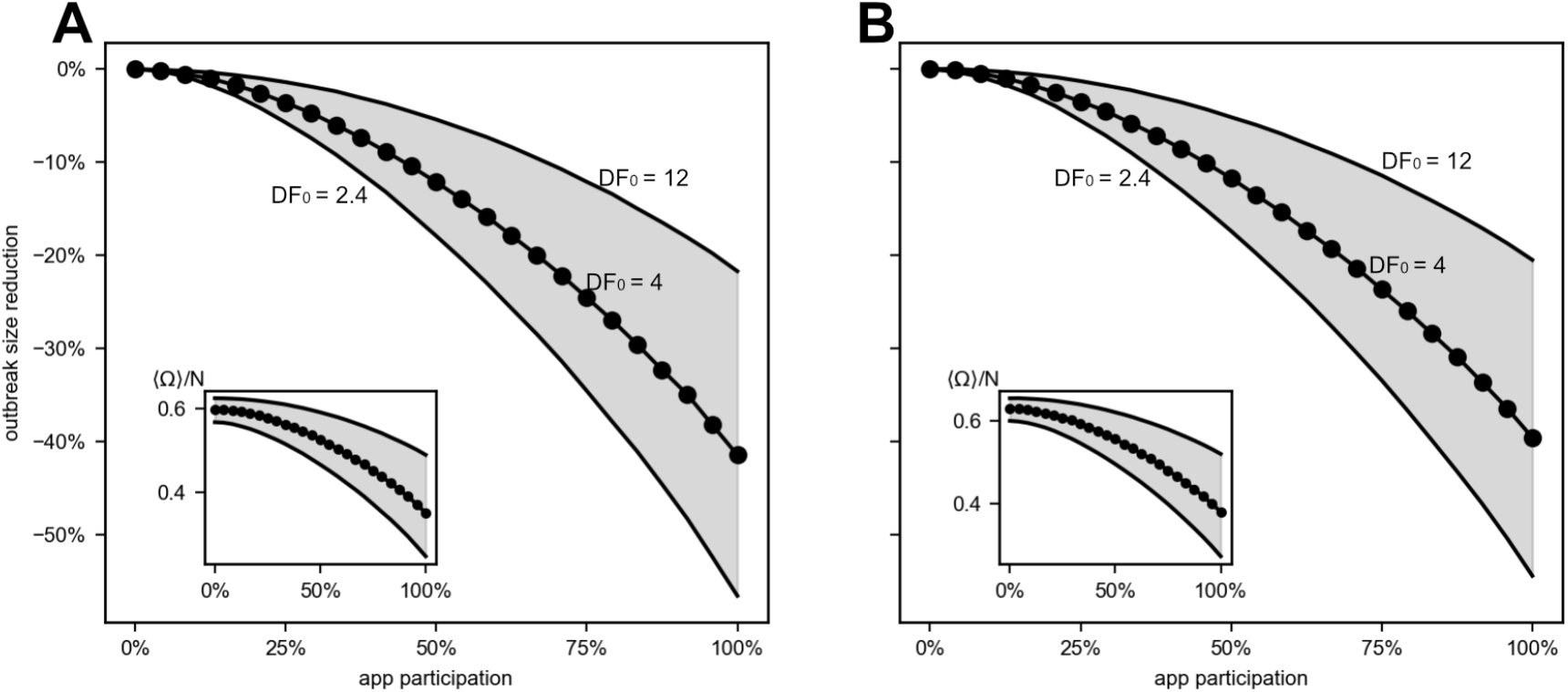
Outbreak size ⟨ Ω ⟩*/N* and relative outbreak size reduction caused by DigCT with *DF*_0_ ∈ {12, 4, 2.4} for increasing app participation *a* in **(A)** assortative small-world network (ordering: ascending) with exponential degree distribution and **(B)** small-world network (ordering: random) with exponential degree distribution. Results between both models do not differ substantially.

### 5. Trajectory of the epidemic influences DigCT’s efficacy

As many countries experienced trajectories where periods of growing case numbers where followed by phases of outbreak suppression (mostly through the introduction of measures that cut contacts in the whole population, i.e. “lockdown” measures), we analyzed how strongly the trajectory of an epidemic influences relative outbreak reduction. We therefore compared this quantity for different trajectories of an epidemic, one where no NPIs other than quarantine and tracing mitigate the disease’s spread and one where the periodic introduction of lockdown measures lead to two consecutive waves. We simulate 100 independent runs on Erdős–Rényi random networks (*N* = 200,000, *k*_0_ = 20, ℛ_0_ = 2.5, *I*_*P*,0_ = 0.001 × *N*).

First, we analyze the trajectory of the otherwise unmitigated epidemic (c.f. Fig. 11A). With rising case numbers, the efficacy of DigCT increases and more cases can be averted per day (as compared to an unmitigated epidemic), both relatively and absolutely (c.f. Fig. 11A.i). Because DigCT has a higher efficacy during the supercritical phase, however, the percentage of averted cases decreases with decreasing prevalence, after a peak has been reached (c.f. Fig. 11A.iii). The DigCT-induced decay of case numbers is slower than in the epidemic where herd immunity was reached without tracing, leading to a negative number of averted cases, reducing the interventions efficacy (c.f. Fig. 11A.iv).

**Figure 11.**
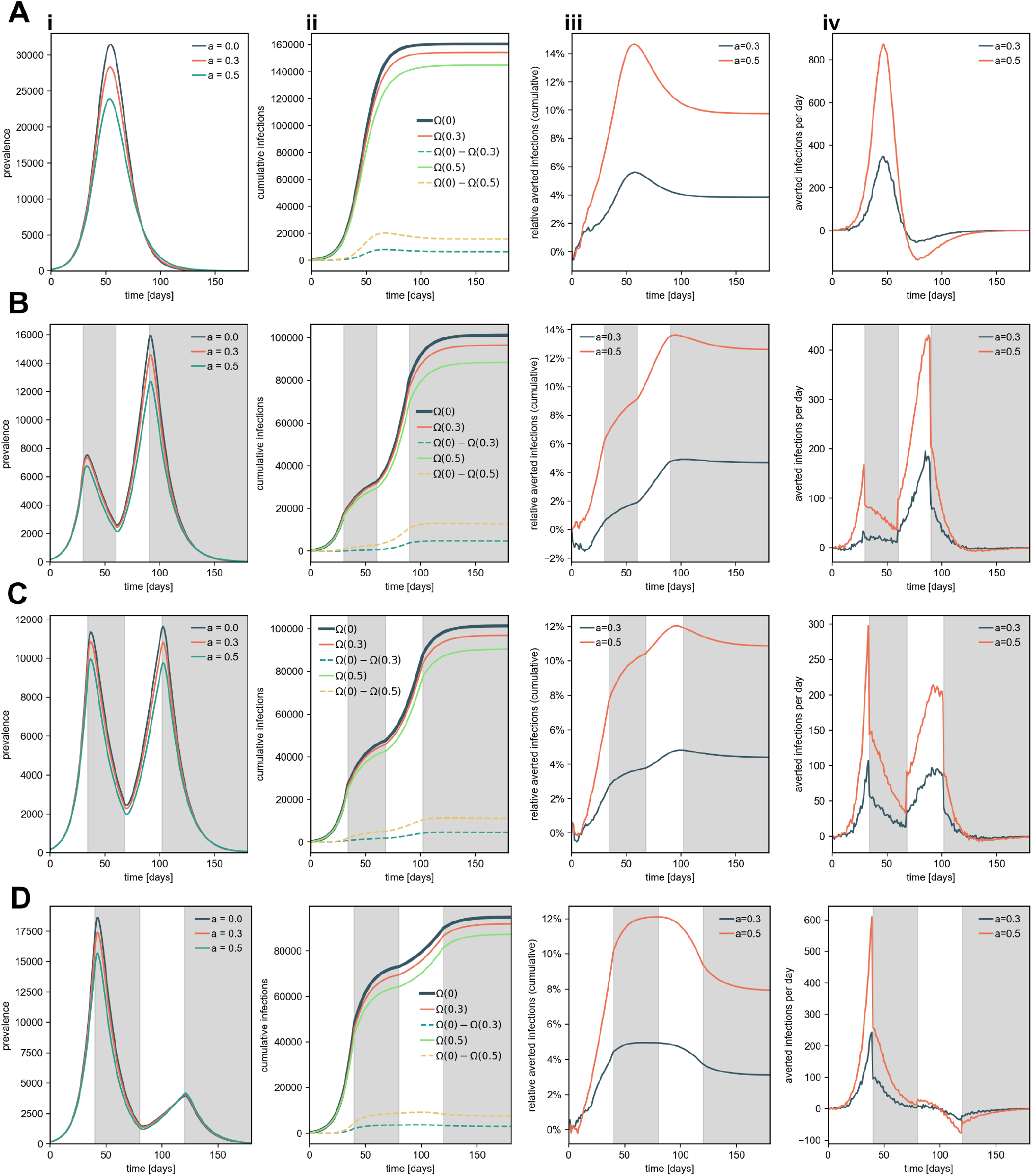
Comparison of **(i)** the prevalence, **(ii)** the cumulative infections Ω_*t*_ (*a*) (and the difference between cumulative infections of “no DigCT” and DigCT-mitigated systems), **(iii)** the relative cumulative averted infections, and **(iv)** the prevented infections per day with *a* ∈ {0%, 30%, 50%} app participation for **(A)** for spread without lockdowns and **(B-D)** for periodically introduced lockdowns where **(B)** *t*_1_ = 30, **(C)** *t*_1_ = 34, **(D)** *t*_1_ = 40.

Simulating multiple other NPIs during the intervention, we introduce “lockdowns” by randomly cutting 60% of links after the first growth period, restoring these links after the first lockdown, and then cutting 50% of links after the second growth phase, keeping this configuration for *t*→ ∞. Lockdown measures were initiated after *t*_1_ days, lifted after *t*_2_ = 2*t*_1_ days, and were reintroduced after *t*_3_ = 3*t*_1_ days. We used *t*_1_ ∈ {30, 34, 40} to compare different trajectories (see Fig. 11): First, a scenario where the second wave is larger than the first wave. Second, a scenario, where the waves are of similar size. In a third scenario, the first wave is much larger than the second. While with *t*_1_ ∈ {30, 34} the efficacy can be increased or maintained, *t*_1_ = 40 leads to lower outbreak size reduction during the epidemic’s subcritical phase. This suggests that the influence of “lockdowns” on DigCT efficacy is highly dependent on an epidemic’s dynamic trajectory and could explain differences of our results to other studies. Note that in these simulations, efficacy only increases strongly when the prevalence reaches high values.

### 6. Influence of random testing during multiple waves

To analyze DigCT’s dependence on testing strategies, we additionally simulated a stripped-down version of our model where we do not differentiate between pre-, a-, and symptomatic infectious individuals (on Erdős-Rényi networks, see Fig. 12). This mimics “random” testing, where the probability to get tested does not depend on specific infection status. For simplicity, we also assume that (i) quarantining an individual leads to immediate notification of their contacts, (ii) every infected contact is immediately quarantined, (iii) no next-generation tracing is possible and (iv) susceptibles will not isolate themselves upon notification. We find a similar effect as described above for the epidemic that is only mitigated by testing: When case numbers rise, the relative number of averted cases increases. After reaching its peak, prevalence decreases slower in the DigCT-controlled system than in the system without tracing, where herd immunity was reached. Hence, the total relative number of averted cases decreases again, reaching a value of 5% for *a* = 0.3 and *q* = 0.3 (c.f. Fig. 12A). However, simulating an epidemic that has been suppressed by other NPIs twice, we find that high values of relative number of averted cases can be reached, that are then not reduced strongly when the outbreak is contained by a harsher reduction of the growth rate trough other interventions (see SI Fig. 12B). From this, we conclude that the total efficacy of DigCT can be kept higher when a larger outbreak is suppressed by the introduction of harsher lockdown measures (in combination with randomized testing).

**Figure 12.**
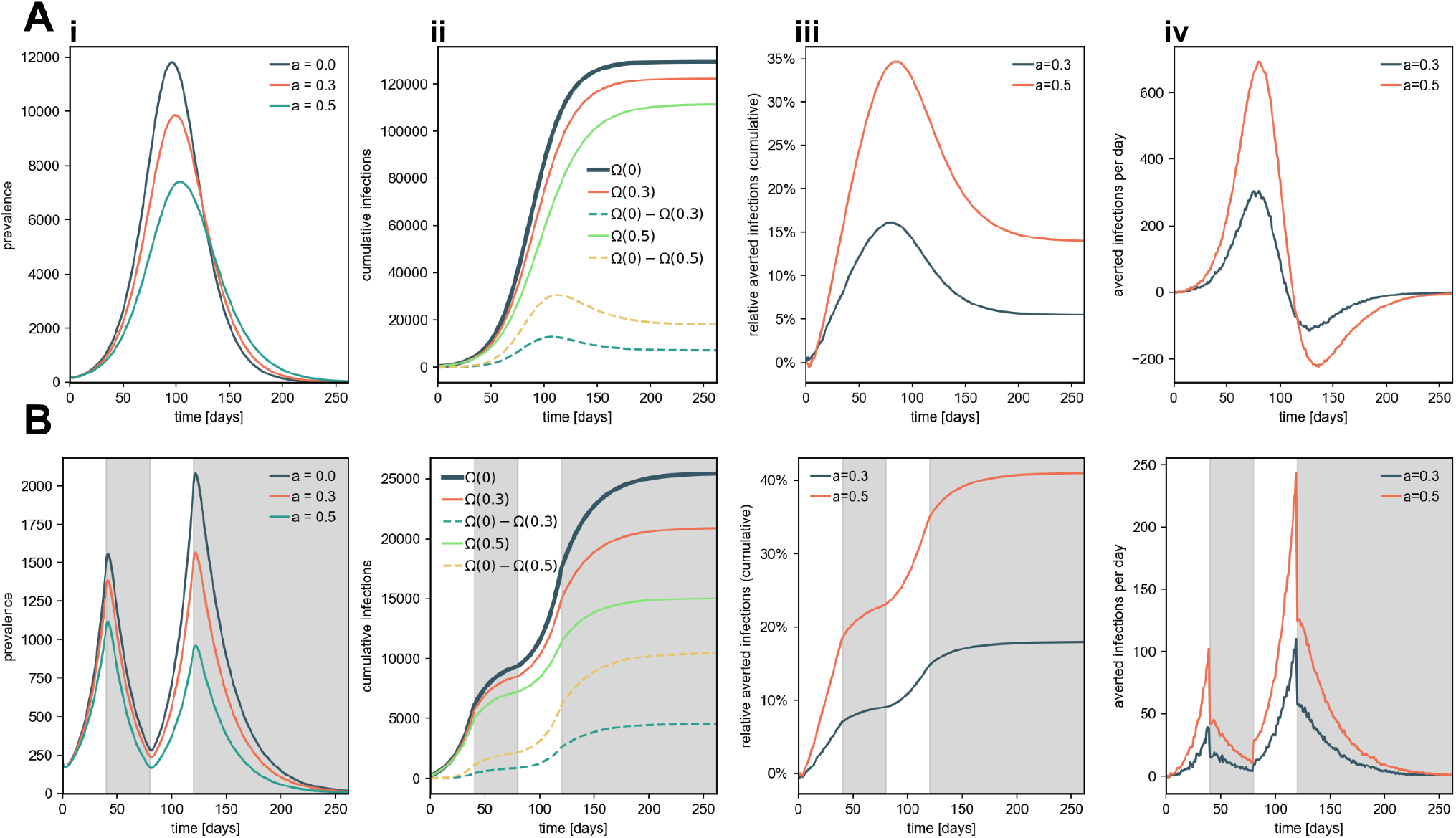
A stripped-down version of our model where we do not differentiate between pre-, a-, and symptomatic infectious individuals. This mimics “random” testing, where the probability to get tested does not depend on specific infection status. We simulate epidemics that are **(A)** only mitigated by quarantine and DigCT as well as **(B)** epidemics that are forced into two waves by other, abstract NPIs (lockdowns, for instance). We show **(i)** the prevalence, **(ii)** the cumulative infections Ω(*t*) (and the difference between cumulative infections of “no DigCT” and DigCT-mitigated systems), **(iii)** relative cumulative averted infections, and **(iv)** averted infections per day with *a* ∈ {0%, 30%, 50%} app participation. While for the (A.iii) otherwise unmitigated disease, DigCT efficacy increases first and decreases afterwards, (B.iii) efficacy monotonically increases in the two-wave system. This illustrates both the influence of the specific trajectory of the epidemic as well as randomized testing. All curves are averages over 100 runs for each simulation. We exclusively simulated on Erdős–Rényi networks with *N* = 200,000 nodes and mean degree *k*_0_ = 20.

